# Mifamurtide for Osteosarcoma in the United States: A Single-Center Real-World Experience

**DOI:** 10.64898/2026.10.04.26364693

**Authors:** Emily K Slotkin, Audrey Mauguen, Jane Roach, Filemon S Dela Cruz, Julia L Glade Bender, Michael V Ortiz, Jovana Pavisic, Olayode O Babatunde, Lauren B Banks, Ping Chi, Sandra P D’Angelo, Mark A Dickson, Mrinal M Gounder, Ciara M Kelly, Mary Lou Keohan, Robert G Maki, Sujana Movva, William D Tap, Laney Berman, Kleoniki Diamantis, Jessie Hennessy, Courtney Henelley, Paulette Kelly, Talia Sauerhaft, Jamie Voccola, Damon R Reed, Leonard H Wexler, Paul A Meyers

## Abstract

**Purpose:** Mifamurtide (liposomal muramyl tripeptide phosphatidylethanolamine [L-MTP-PE]) is approved in many countries for osteosarcoma but remains unavailable for routine use in the United States (US). We describe the feasibility and outcomes of patients with osteosarcoma treated with mifamurtide at Memorial Sloan Kettering Cancer Center (MSK) through expanded access (EAP) and compassionate use (SPU) pathways.

**Methods:** Medical records were retrospectively reviewed for patients with completely resected, high-grade osteosarcoma treated with mifamurtide at MSK from 2016–2026 through EAP or SPU protocols. Mifamurtide 2 mg/m² was administered intravenously twice weekly for 12 weeks, followed by weekly administration for 24 weeks. Outcomes were evaluated by treatment setting and, for upfront therapy, metastatic status and histologic necrosis.

**Results:** Fifty-seven patients received mifamurtide (median age, 14 years; range, 2-60); 72% had localized and 28% metastatic disease at diagnosis. No serious adverse events were considered possibly or probably related to mifamurtide, and no new grade 3-4 safety signals emerged. Five patients with localized disease and ≥90% necrosis at resection had no events, with 100% progression-free survival (PFS) and overall survival (OS) at median follow-up of 29 months. Among 22 patients with localized disease and <90% necrosis treated upfront, PFS was 82%, 66%, and 59% at 12, 24, and 48 months, respectively; OS was 95% at 12, 24, and 60 months. Among 8 patients with metastatic disease treated upfront, PFS was 73%, 58%, and 39% at 12, 24, and 36 months, respectively; OS was 100%, 86%, and 86%. Among 22 patients treated after relapse, PFS was 34%, 23%, and 17% at 12, 24, and 36 months, respectively; OS was 91%, 70%, and 35% at 12, 24, and 60 months.

**Conclusions:** In this real-world US cohort, mifamurtide was feasible and safe to administer. Despite limitations of the EAP/SPU setting, single-institution administration, and patient heterogeneity, overall survival outcomes were encouraging.

## Introduction

Osteosarcoma is the most common primary malignant tumor of bone, arising predominantly in children, adolescents, and young adults, with a peak incidence that coincides with the pubertal growth spurt and a smaller second peak in later adult life^1–3^. High-intensity chemotherapy with cisplatin, doxorubicin and high-dose methotrexate (MAP) and complete surgical resection of radiographically visible sites of disease when feasible remains the global standard of care^4–6^. Patients with radiographically localized and resectable disease achieve a five-year overall survival ranging from ∼70-80%^7–10^, whereas those presenting with metastatic disease or relapse fare considerably worse with post-relapse five-year survival less than 30%^11,12^. Attempts to improve upon the MAP regimen through intensification or the addition of new agents have been unsuccessful^6,7,13^, and in the United States no new systemic agent has modified the standard of care in many decades^14–16^ underscoring a persistent and widely recognized unmet need^17,18^.

Mifamurtide (liposomal muramyl tripeptide phosphatidylethanolamine (L-MTP-PE)), a synthetic analogue of a bacterial cell-wall peptidoglycan that activates monocytes and macrophages to a tumoricidal state and triggers a cascade of proinflammatory cytokines^19^, is approved across Europe and other regions. The Intergroup Study 0133 (CCG-7921, POG-9351) was a prospective, randomized phase III trial in newly diagnosed patients 30 years of age or younger with osteosarcoma conducted from November 1993 to November 1997^5^. The study used a 2×2 factorial design designed to answer whether the addition of either ifosfamide or mifamurtide to the MAP regimen would improve EFS. In an initial manuscript published in 2005, six hundred and seventy-seven osteosarcoma patients were randomized to receive 1 of 4 regimens after definitive surgery: the MAP regimen alone (A-), the MAP regimen with mifamurtide (A+), the MAP regimen with ifosfamide (B-), and the MAP regimen with ifosfamide and mifamurtide (B+). An interaction observed between mifamurtide and ifosfamide prevented statistical analysis using the planned factorial design and estimates of EFS according to each treatment arm were instead provided with the MAP regimen alone (A-) resulting in 3-year EFS of 71%, MAP with mifamurtide (A+) in 3-year EFS of 68%, MAP with ifosfamide (regimen B−) in 61% 3-year EFS, and MAP with ifosfamide and mifamurtide in 78% 3-year EFS. The definitive improvement of EFS with the addition of mifamurtide could not be proven with these initial data. However, a follow up report published in 2008 with the benefit of more prolonged follow up eliminated the interaction between ifosfamide and mifamurtide and showed no benefit with the addition of ifosfamide to the MAP regimen, but did find a trend toward improved EFS and statistically significant improvement in 6-year overall survival (OS) from 70% to 78%^20^. These data facilitated approval for use of mifamurtide for the treatment of osteosarcoma in many areas internationally, but not in the United States. Since that time the use of mifamurtide in the United States has been irregular and sporadic, with motivated providers and patients utilizing EAP and SPU pathways when feasible^21^.

Evidence of the real-world effectiveness of mifamurtide outside this pivotal trial remains limited and is drawn largely from small single-institution series^22–25^.The role of the agent in radiographically localized disease continues to be debated, and a recent analysis of two prospective European trials in P-glycoprotein– positive patients reignited that discussion^26,27^. Real-world cohorts that capture the heterogeneous populations treated under expanded access, spanning pediatric, adolescent, and adult patients with both newly diagnosed and relapsed disease, are therefore of particular value.

We report a single-center retrospective cohort of 57 patients from the United States with high-grade osteosarcoma treated with mifamurtide, 49 of which were treated on an institutional EAP (NCT04571229) and 8 of which were treated via SPU protocols over a 10-year period. The objective was to describe PFS and OS outcomes of this real-world cohort and to document the feasibility and tolerability of treatment delivery of mifamurtide in routine clinical practice.

## Methods

### Study Participants and Data Collection

Records from patients with completely resected high grade osteosarcoma who received mifamurtide via EAP or SPU at Memorial Sloan Kettering Cancer Center (MSK) from 2016 to 2026 were reviewed. The retrospective record review, the EAP (NCT04571229), and each SPU were conducted in accordance with the Declaration of Helsinki, approved by the MSK Institutional Review Board (IRB) and the US Food and Drug Administration (FDA). Written informed consent for treatment was obtained from every patient or legal guardian, and assent was obtained from every patient from 7 to 17 years of age. Information was de-identified and clinical variables were abstracted from electronic medical records.

EAP and SPU participation required a diagnosis of high-grade osteosarcoma deemed completed resected by the treating oncologist and acceptable standard organ function (Protocol, online only).

### Treatment

Mifamurtide was administered by intravenous (IV) infusion at a dose of 2 mg/m^2^ twice weekly for the first 12 weeks and then once weekly for 24 weeks, for a planned total of 48 doses delivered over 36 weeks (Figure 1). Mifamurtide was given as monotherapy or in addition to standard chemotherapy regimens (e.g., MAP, ifosfamide/etoposide) at the discretion of the investigators (Protocol, online only). Premedication with medications such as acetaminophen and/or other supportive measures were used as clinically appropriate, including the use of meperidine 0.5 mg/kg (up to a maximum of 25 mg IV) for infusion associated chills or rigors. Grade 3 and 4 adverse events and serious adverse events were collected. Hospitalization for the management of febrile neutropenia related to concomitant chemotherapy administration was not collected as a serious adverse event.

**Figure 1:**
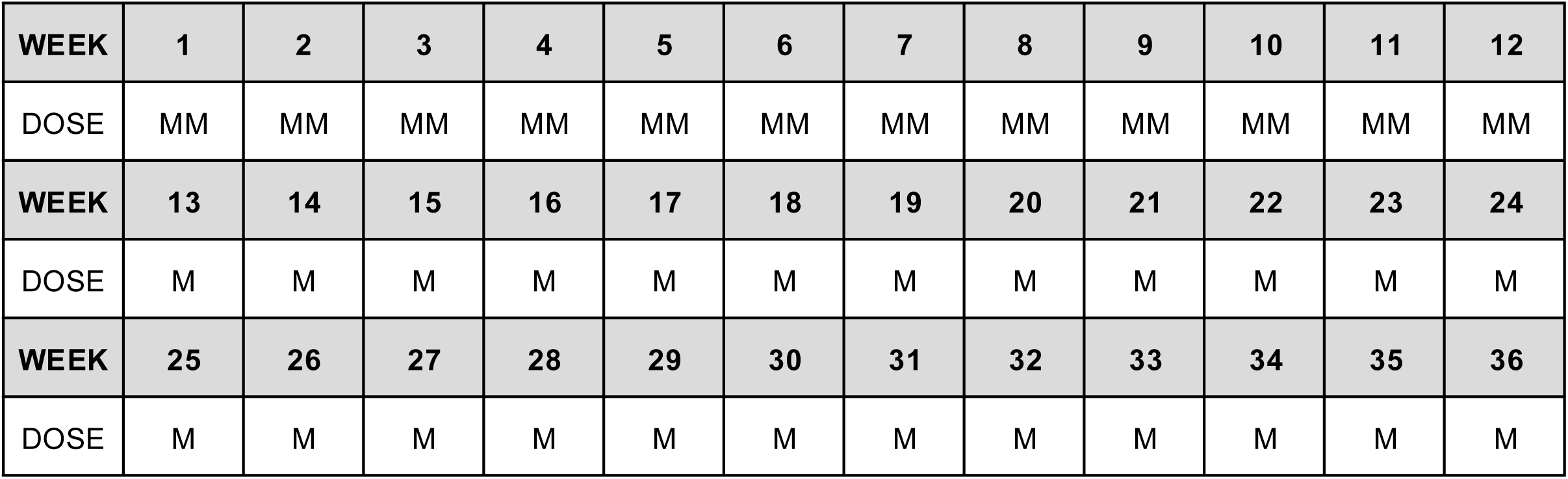
Schema for mifamurtide administration.

### Endpoints

Patients were grouped according to phase of therapy (upfront or relapsed/refractory). PFS was measured from the date of the first mifamurtide dose to the first occurrence of documented disease progression, relapse, or death from any cause, whichever came first. OS was measured from the first mifamurtide dose to death from any cause. Patients who had not experienced any event of interest were censored at the date of last contact using a data cutoff of August 2026. Histological necrosis was recorded from definitive resection of the presumed primary site of disease regardless of the phase of therapy in which mifamurtide was used. Data was stratified according to histological necrosis for patients receiving mifamurtide in the upfront treatment setting.

### Statistical Analysis

Baseline characteristics were summarized using medians and ranges for continuous variables and counts with percentages for categorical variables. Time to event endpoints were estimated by the Kaplan-Meier estimator, and survival probabilities are reported together with their 95% confidence intervals. Median follow-up times were estimated using the reverse Kaplan-Meier. Outcomes were estimated separately for the upfront and the relapsed or refractory cohorts.

## Results

### Patient and Tumor Characteristics

Fifty-seven patients with completely resected high-grade osteosarcoma who received mifamurtide on either the EAP or an SPU from 2016 to 2026 at our institution were included in the analysis (Table 1). The median age at diagnosis was 14 years (range 2-60 years) with a slight male predominance (34 patients, 60%). Primary tumor in an extremity was most common (49 patients, 86%). Disease at diagnosis was radiographically localized in 41 patients (72%) and metastatic in 16 patients (28%) (Table 1). Histologic necrosis with upfront resection (regardless of phase of therapy during which mifamurtide was given) was available for all but two patients. Twenty patients (36%) had less than 50% necrosis, 20 patients (36%) had 50-89% necrosis, 13 patients (24%) had necrosis of 90-99%, and 2 patients (4%) had necrosis of 100% (Table 1, Supplementary Table 1).

**Table 1.**

| Characteristics | n (%); total n=57 |
| --- | --- |
| **Sex** |  |
| Female | 23 (40) |
| Male | 34 (60) |
| **Age at Diagnosis** (median [range]) | 14 [2 – 60] |
| **Stage at Diagnosis** |  |
| Localized | 41 (72) |
| Metastatic | 16 (28) |
| **Primary Site at Diagnosis** |  |
| Extremity | 49 (86) |
| Other | 4 (7) |
| Pelvis | 4 (7) |
| **Histologic Necrosis at Upfront Resection** |  |
| < 50% | 20 (36) |
| 50-89% | 20 (36) |
| 90-99% | 13 (24) |
| 100% | 2 (4) |
| unknown | 2 |
| **Phase of Therapy During Which MTP Received** |  |
| Upfront, in first remission | 35 (61) |
| Relapse in radiographic remission | 22 (39) |

### Treatment Administration

No patient experienced a serious adverse event (defined as death, a life-threatening adverse event, an adverse event that results in inpatient hospitalization or prolongation of existing hospitalization, a persistent or significant incapacity or substantial disruption of the ability to conduct normal life functions, a congenital anomaly/birth defect, or other Important Medical Event (IME) that may not result in death, be life threatening, or require hospitalization but may jeopardize the patient or participant and may require medical or surgical intervention) possibly or probably related to mifamurtide administration. Treatment was delivered in the ambulatory setting unless inpatient care was otherwise clinically indicated. The safety and tolerability of the use of mifamurtide as both monotherapy and in combination with chemotherapy is well established^5,20,25,27–29^, and no new grade 3 or 4 adverse event safety signals were identified when compared with these prior published experiences. Data regarding the frequency of grade 1 or 2 adverse events, including the frequency of chills/rigors or fevers associated with mifamurtide administration, are not available given the EAP or SPU setting of administration.

### Outcomes

Only 5 patients had radiographically localized disease, received mifamurtide in the upfront treatment setting, and had ≥90% necrosis. At the time of data cutoff these patients had been followed for 6, 12, 29, 56 and 81 months (median 29 months) respectively and none of them had experienced an event or death conferring both 100% PFS and OS.

Twenty-two patients had radiographically localized disease, received mifamurtide in the upfront treatment setting and had <90% necrosis (Figure 2). PFS for these patients at 12 months was 82% (95% CI 67-100%), at 24 months was 66% (95% CI 48-90%), and at 48 months was 59% (95% CI 40-86%) and OS was 95% at 12, 24 and 60 months (95% CI 86-100%) (Figure 2A-B) with median follow-up of 43 months.

**Figure 2:**
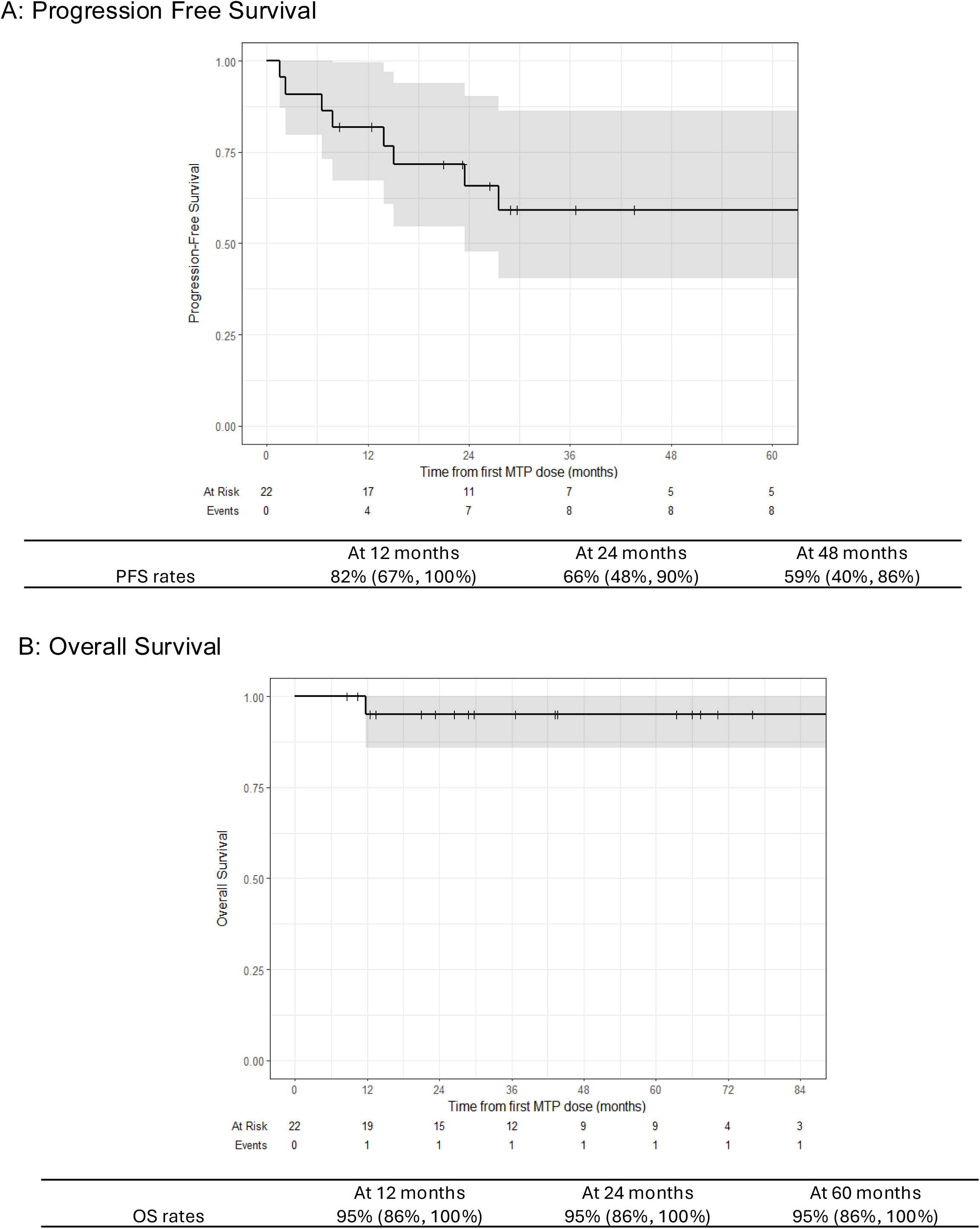
Outcomes for patients with radiographically localized and completed resected osteosarcoma with necrosis <90% who received mifamurtide in the upfront treatment setting.

Eight patients had metastatic disease and received mifamurtide in the upfront treatment setting. PFS for these patients at 12 months was 73% (95% CI 47-100%), at 24 months was 58% (95% CI 31-100%), and at 36 months was 39% (95% CI 14-100%) and OS was 100% at 12 months, 86% at 24 months (95% CI 63-100%), and 86% at 36 months (95% CI 63-100%) (Figure 3A-B) with median follow-up of 38 months.

**Figure 3:**
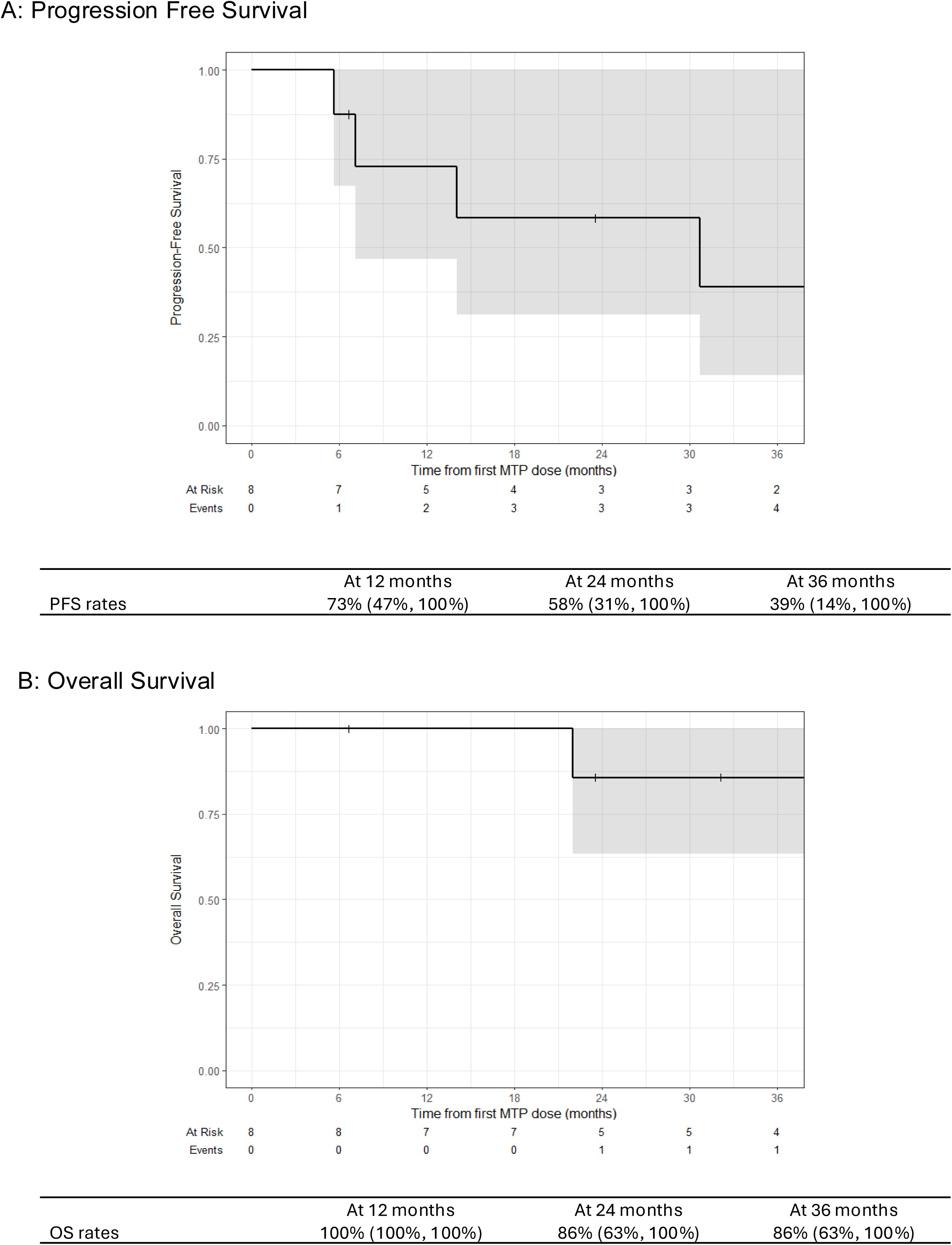
Outcomes for patients metastatic and completed resected osteosarcoma who received mifamurtide in the upfront treatment setting.

Twenty-two patients received mifamurtide in the relapsed/refractory but completely resected setting. PFS for these patients was 34% at 12 months (95% CI 19-62%), 23% at 24 months (95% CI 10-52%), and 17% at 36 months (95% CI 6-46%) and OS was 91% at 12 months (95% CI 80-100%), 70% at 24 months (95% CI 52-94%), and 35% at 60 months (95% CI 17-71%) (Figure 4A-B) with median follow-up 67 months.

**Figure 4:**
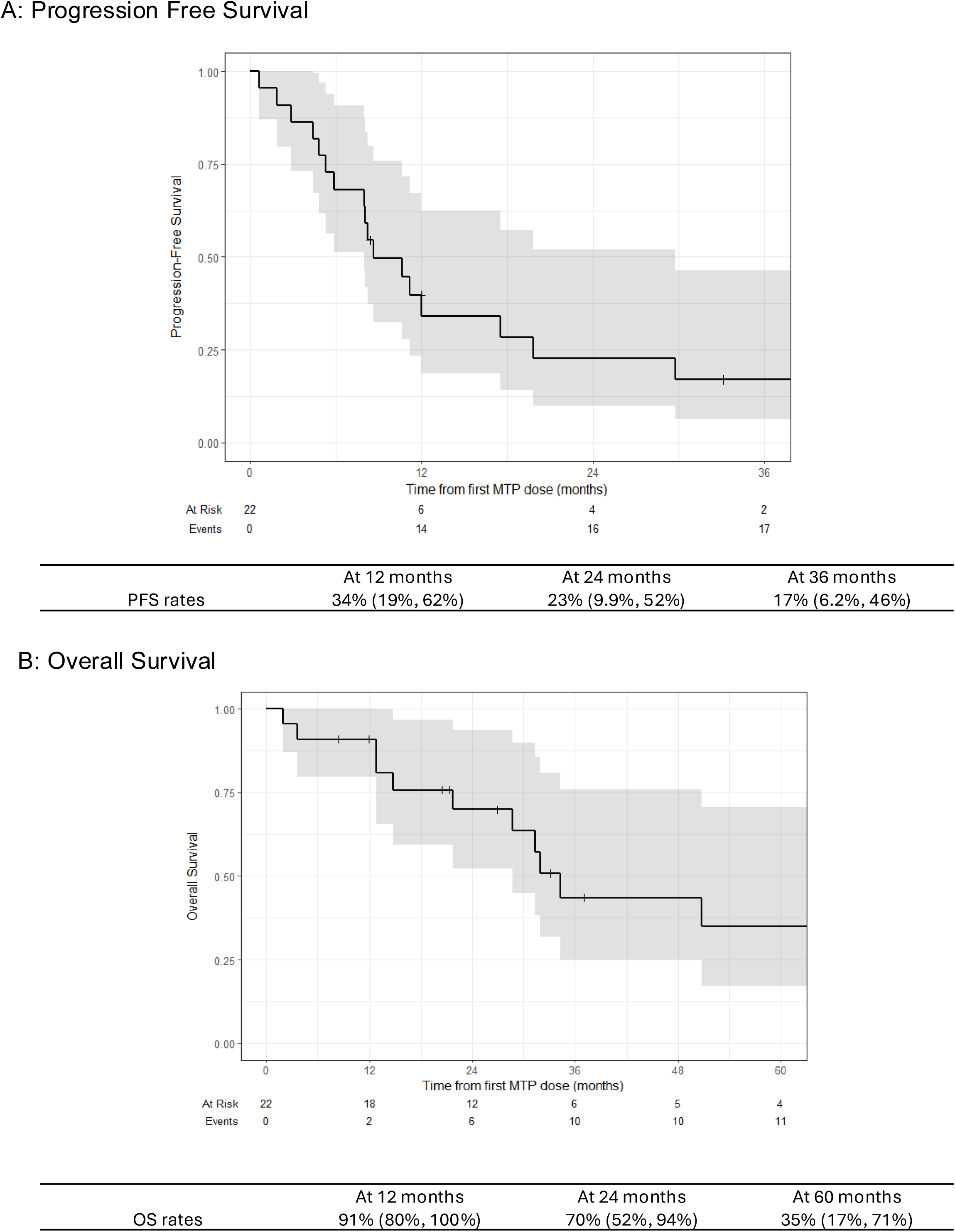
Outcomes for patients who received mifamurtide in the relapsed and completely resected setting.

## Discussion

In this real-world experience of 57 patients with high-grade osteosarcoma, mifamurtide was safely delivered in the outpatient setting across the full clinical spectrum encountered in practice using expanded access (NCT04571229) and single patient use protocol approaches. Although this experience is notably limited by its retrospective nature, single-center administration, and modest patient numbers, these data add to the growing body of real-world experiences^22–25^ using mifamurtide in the upfront treatment setting which achieve overall survival outcomes more favorable than those without the use of mifamurtide^6,30^. In the relapsed or refractory setting, PFS rates were predictably low reflecting the significant unmet need of this patient population.

Continued interest in mifamurtide reflects its non-toxic nature and the scarcity of effective systemic agents in osteosarcoma. Attempts to improve outcomes with chemotherapy intensification^6^, interferon^13^, checkpoint inhibition^31^ and numerous additional novel agents^14–16,32–34^ have failed to change the standard of care. In this context mifamurtide represents a tolerable immunomodulatory agent with an existing survival signal and decades of clinical experience, and therefore retains a rational place in the treatment of patients with osteosarcoma.

In October 2025, the FDA co-hosted a workshop in collaboration with the Osteosarcoma Institute (OSI), bringing together more than 370 academic investigators, clinical trialists, regulatory scientists, and patient advocacy representatives to address major scientific and clinical barriers to drug development in osteosarcoma^35^. The workshop reinforced that progress requires not only the rapid identification of promising agents and attention to the molecular heterogeneity of osteosarcoma, but also innovative trial designs and endpoints, because large, randomized phase 3 trials are difficult to conduct in such a rare disease^18,35^. In this context, real-world datasets such as the one presented here are constructive because they provide signals from the heterogeneous populations treated in practice and may contribute to forward momentum in an otherwise relatively bleak drug development landscape at this time.

In this real-world and retrospective cohort spanning the full age and risk spectrum of high-grade osteosarcoma, mifamurtide was associated with durable disease control and excellent overall survival when incorporated into upfront multimodal therapy. Treatment was well tolerated, with no new safety concerns identified. As the osteosarcoma community intensifies efforts to overcome a four-decade survival plateau, mifamurtide remains a pragmatic and biologically grounded option whose real-world effectiveness warrants continued, systematic documentation.

## Data Availability

All data produced in the present study are available upon reasonable request to the authors.

---

**Supplementary Table 1.**

| Patient Number | Primary Site | % necrosis with upfront primary site resection | PFS (months) | OS (months) |
| --- | --- | --- | --- | --- |
| **Radiographically Localized, Upfront Mifamurtide, ≥ 90% necrosis (total n = 5)** |  |  |  |  |
| 1 | extremity | 90-95% | 6 | 6 |
| 2 | extremity | 99% | 81 | 81 |
| 3 | extremity | 97% | 29 | 29 |
| 4 | extremity | 95% | 56 | 56 |
| 5 | pelvis | 90% | 12 | 12 |
| **Radiographically Localized, Upfront Mifamurtide, <90% necrosis (total n = 22)** |  |  |  |  |
| 6 | extremity | 80-90% | 12 | 12 |
| 7 | extremity | 80-85% | 29 | 29 |
| 8 | extremity | 50-55% | 23* | 98 |
| 9 | extremity | 10-15% | 67 | 67 |
| 10 | extremity | <5% | 69* | 70 |
| 11 | extremity | 10% | 7* | 66 |
| 12 | extremity | 85% | 13* | 118 |
| 13 | extremity | 80% | 27* | 43 |
| 14 | extremity | 80% | 36 | 36 |
| 15 | extremity | 80% | 26 | 26 |
| 16 | extremity | 70% | 20 | 20 |
| 17 | extremity | 65% | 23 | 23 |
| 18 | extremity | 60% | 1* | 13 |
| 19 | extremity | 60% | 43 | 43 |
| 20 | extremity | 60% | 28 | 28 |
| 21 | extremity | 40% | 10 | 10 |
| 22 | extremity | 15% | 15* | 96* |
| 23 | other | 10% | 67 | 67 |
| 24 | extremity | 10% | 76 | 76 |
| 25 | pelvis | 10% | 2* | 11* |
| 26 | extremity | 10% | 8 | 8 |
| 27 | extremity | 5% | 63 | 63 |
| **Metastatic Disease, Upfront Mifamurtide (total n = 8)** |  |  |  |  |
| 28 | extremity | >95% | 5* | 32 |
| 29 | extremity | 100% | 6 | 6 |
| 30 | extremity | 99% | 41 | 41 |
| 31 | extremity | 70% | 30* | 41 |
| 32 | extremity | 60% | 23 | 23 |
| 33 | extremity | 45% | 7* | 28* |
| 34 | extremity | 20% | 38 | 38 |
| 35 | extremity | 15% | 14* | 68 |
| **Relapsed or Refractory Disease (n = 22)** |  |  |  |  |
| 36 | other | unknown | 68 | 68 |
| 37 | other | unknown | 0* | 1* |
| 38 | extremity | 90-95% | 8* | 20 |
| 39 | extremity | 60-70% | 7* | 31* |
| 40 | extremity | >90% | 5* | 12* |
| 41 | extremity | <5% | 11 | 11 |
| 42 | extremity | 100% | 5* | 31* |
| 43 | extremity | 98% | 8* | 21 |
| 44 | extremity | 95% | 11* | 12* |
| 45 | extremity | 94% | 19* | 50* |
| 46 | extremity | 90% | 4* | 34* |
| 47 | extremity | 85% | 8 | 8 |
| 48 | extremity | 80% | 33 | 33 |
| 49 | extremity | 70% | 69 | 69 |
| 50 | extremity | 60% | 12* | 26 |
| 51 | extremity | 50% | 2* | 36 |
| 52 | extremity | 45% | 1* | 3* |
| 53 | extremity | 35% | 4* | 14* |
| 54 | extremity | 30% | 29* | 67 |
| 55 | pelvis | 15% | 10* | 70 |
| 56 | other | 10% | 17* | 28* |
| 57 | pelvis | 0% | 8* | 21* |

