## Supplementary material for "Mifamurtide for Osteosarcoma in the United States: A Single-Center Real-World Experience": Figure Legends

**Table 1**: characteristics of patients with completely resected high-grade osteosarcoma who received mifamurtide at Memorial Sloan Kettering Cancer Center (MSK) via expanded access (EAP, NCT04571229) or single patient use (SPU) protocols

**Figure 1:** schema of mifamurtide administration

**Figure 2:** outcomes for patients with radiographically localized and completed resected osteosarcoma with necrosis <90% who received mifamurtide in the upfront treatment setting; 2A = progression free survival; 2B = overall survival

**Figure 3:** outcomes for patients with metastatic and completed resected osteosarcoma who received mifamurtide in the upfront treatment setting; 3A = progression free survival; 3B = overall survival

**Figure 4:** outcomes for patients with completed resected osteosarcoma who received mifamurtide in the relapsed or refractory treatment setting; 4A = progression free survival; 4B = overall survival

**Supplementary Table 1**: individual necrosis, progression free survival and overall survival data for each patient receiving mifamurtide
